# Genetic support for an adverse effect of calcium channel blockers on the human thoracic aorta

**DOI:** 10.64898/2026.10.02.26364570

**Authors:** Julia Horjus, Sean J. Jurgens, Connie R. Bezzina, Nimrat Grewal

## Abstract

Despite the widespread use of L-type calcium channel blockers (CCBs), recent observational and murine functional work has suggested a potential adverse effect of this medication class on the aorta. Here, we apply a Mendelian randomization (MR) framework to assess the potential causal role of *CACNA1C* - the principal target of L-type CCBs in the human aorta - on thoracic aortic aneurysm and dissection (TAA/D). Genetic instrumental variables were derived from aortic expression quantitative trait locus data, and combined with large-scale genome-wide association data of TAA/D and ascending aortic diameter. Mirroring the on-target vascular effects of L-type CCBs, higher genetically predicted *CACNA1C* expression was linked to higher blood pressure, and to higher risks of hypertension and coronary artery disease. In contrast, higher predicted aortic expression of *CACNA1C* was linked to lower risk of TAA/D (β=-0.18, *P*=0.005) and to a smaller ascending aortic diameter in the general population (β=-0.08, *P*=0.005). While we caution against overinterpretation of our MR results in isolation, the convergence with observational and functional data calls for dedicated clinical trials to assess the safety of CCBs in the context of aortic disease.

## Main

Thoracic aortic aneurysm represents the pathological and irreversible enlargement of the thoracic aorta, which may lead to dissection - an acute tearing of the intimal layer of the aortic wall. Thoracic aortic dissection is a life-threatening medical emergency with a high mortality rate^1^. While surgical intervention is the gold-standard for specific types of dissection, blood pressure control remains fundamental in the management of both acute dissections and thoracic aneurysms^2,3^.

Indeed, hypertension has long been recognized as an independent risk factor for thoracic aortic aneurysms and dissections (TAA/D)^4,5^. In clinical practice, antihypertensive drugs - including β-blockers, angiotensin II receptor blockers, and L-type calcium channel blockers (CCBs) - are commonly prescribed in TAA/D. These same classes of medication are also frequently prescribed in patients with other cardiovascular diseases or with elevated cardiovascular risk profiles.

Despite their widespread use, recent work has suggested that L-type CCBs may confer harm to the aorta (**Figure 1A**). Ma et al.^6^ found that CCB users experience higher risk of TAA/D compared to hypertensive patients not receiving antihypertensive treatment. Furthermore, in the same study, dissection patients who underwent endovascular repair showed more limited aortic regression with CCBs as compared to other antihypertensives. Murine functional work also supports a direct effect of CCBs on aortic stiffness and aneurysm formation^6^. Nevertheless, all current human evidence is based on observational data, which is highly liable to confounding. Therefore, it remains unclear whether CCBs are causally linked to TAA/D risk in humans.

**Figure 1.**
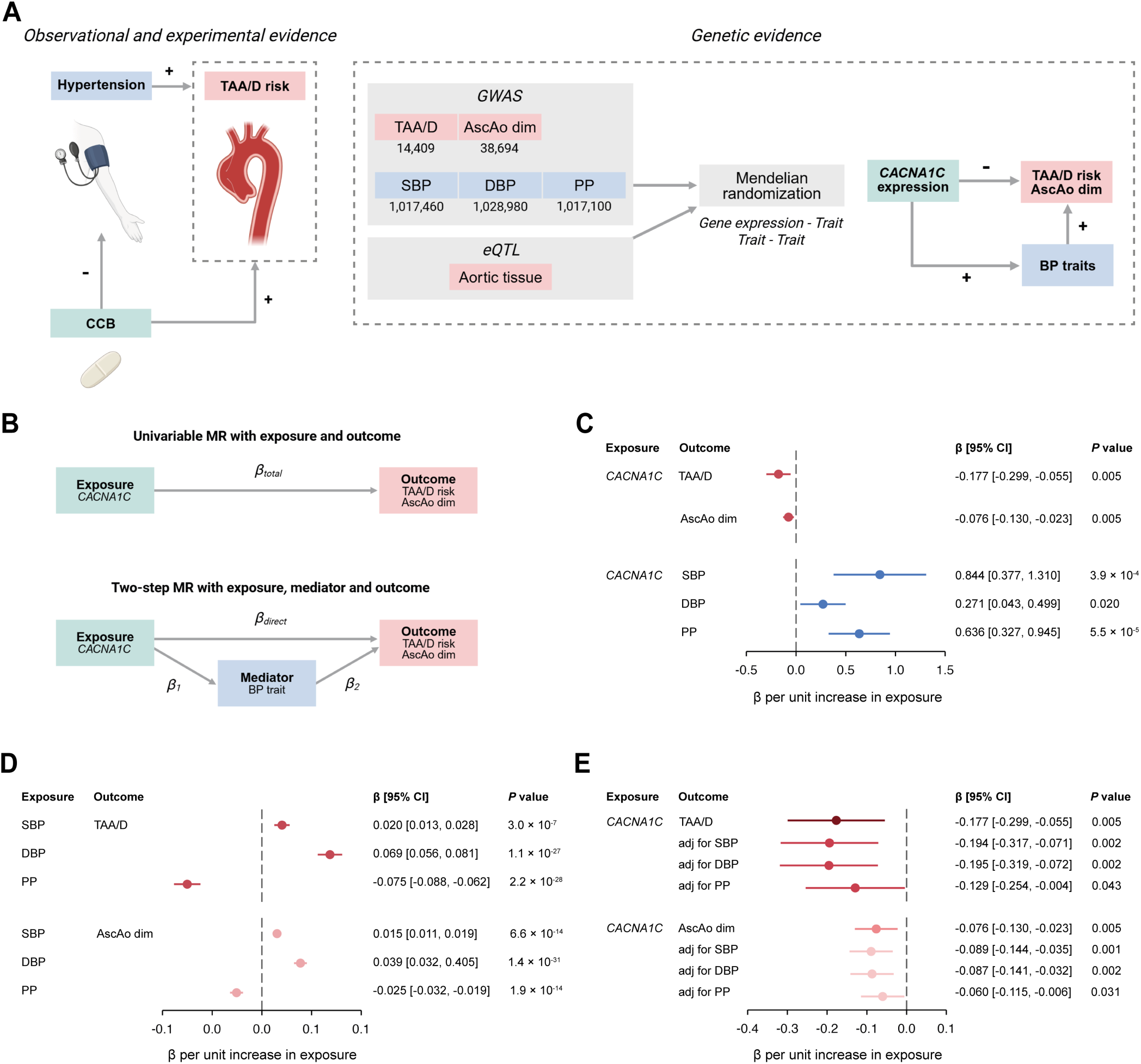
Genetic evidence from *CACNA1C* aortic expression supports adverse effects of calcium channel blocker use in thoracic aortic disease. A, Study overview. Previous observational and experimental evidence suggests adverse effects of CCB use on TAA/D risk (left panel). These effects are recapitulated with human genetic evidence, by performing MR analyses of aortic *CACNA1C* expression on both TAA/D risk and ascending aortic diameter, while accounting for blood pressure traits (right panel). B, MR analyses to assess the effect of aortic *CACNA1C* expression on TAA/D risk and ascending aortic diameter. A potential mediation effect of blood pressure traits is accounted for using a two-step MR approach. C, Forest plot showing the results for univariable MR of aortic *CACNA1C* expression on both aortic outcomes and blood pressure traits. D, Forest plot showing the results of univariable MR for the effect of blood pressure traits on both aortic outcomes. E, Forest plot showing the results of two-step MR for the effect of aortic *CACNA1C* expression on aortic outcomes, adjusting for a potential mediation effect of blood pressure on both TAA/D risk and ascending aortic diameter.

Here, we aimed to find supportive evidence for a causal role of CCBs in TAA/D risk using human genetic evidence (**Figure 1A**; **Methods**). Briefly, in a Mendelian randomization framework, we used genetic variants as proxies (instrumental variables) for aortic expression of *CACNA1C* – which encodes the main pore-forming subunit of the arterial-expressed calcium channel and which represents the main genetic target of antihypertensive L-type CCBs^7,8^. Of note, while other pore-forming subunits (*CACNA1D, CACNA1S, CACNA1F*) also represent targets of CCBs, these subunits are barely expressed in the human thoracic aorta^9^.

For our Mendelian randomization, we used a multi-SNP instrument model based on a published sparse expression model for *CACNA1C* in aorta^10^, built from expression quantitative trait loci (QTL) data in GTEx v8^11^ (**Supplementary Table 1**; **Methods**). The instruments for *CACNA1C* expression were integrated with large genome-wide association (GWAS) data for TAA/D^12^ (N=14,409 cases and N=1,210,227 controls; **Supplementary Table 2**) and a GWAS on ascending aortic diameter in the general population^9^ (N=38,694 participants). We similarly used Mendelian randomization to validate and explore other causal links between *CACNA1C* expression, blood pressure traits, and TAA/D (**Figure 1AB**).

We first aimed to validate our *CACNA1C* expression model in the context of known L-type CCB effects. As expected, Mendelian randomization linked higher *CACNA1C* expression in aorta to higher systolic blood pressure (β=0.84; 95% CI [0.38, 1.31], *P*=3.9×10^-4^), higher diastolic blood pressure (β=0.27; 95% CI [0.04, 0.50], *P*=0.02), and higher pulse pressure (β=0.64; 95% CI [0.33, 0.95], *P*=5.5×10^-5^) (**Figure 1C**; **Supplementary Table 3**). These findings mirror the known clinical effects of L-type CCBs - *CACNA1C* inhibitors - on blood pressure traits^13,14^. Consistently, Mendelian randomization linked higher *CACNA1C* expression to higher risk for diagnoses of hypertension (β=0.14; 95% CI [0.07, 0.21], *P*=6.0×10^-5^) and coronary artery disease (β=0.08; 95% CI [0.04, 0.15], *P*=0.038) (**Supplementary Table 4**) – conditions for which L-type CCBs are commonly prescribed^15,16^.

Next, we aimed to replicate the established role of blood pressure on risk of TAA/D within a Mendelian randomization framework, using the weighted median method which is robust to outliers^17^. Indeed, as expected, higher systolic and diastolic blood pressure was significantly linked to higher TAA/D risk and larger ascending aortic diameter (**Figure 1D**; **Supplementary Table 5**), whereas pulse pressure showed the opposite directional effect. These findings are consistent with previous observational and Mendelian randomization reports^18,19^.

We then aimed to estimate the potential causal effect of aortic *CACNA1C* expression on TAA/D and thoracic aortic dimensions. Mendelian randomization linked higher aortic expression of *CACNA1C* to lower risk of TAA/D (β=-0.18, 95% CI [−0.30, - 0.05], *P*=0.005), and to a smaller ascending thoracic aortic diameter in the general population (β=-0.08, 95% CI [−0.13, −0.02], *P*=0.005) (**Figure 1C**; **Supplementary Table 3**). Notably, these findings are directionally consistent with the observational evidence presented in Ma et al.^6^, who showed that CCB use was associated with higher TAA/D risk.

In the work by Ma et al.^6^, the authors also reported adverse effects of CCBs on the abdominal aorta. While we indeed saw a consistent signal for our *CACNA1C* instruments using an existing multi-ancestry meta-analysis of abdominal aortic aneurysm (β=-0.02, *P*=0.004), this effect became insignificant in certain sensitivity analyses (**Supplementary Table 6**). Therefore, we caution against overinterpretation of these results at this stage.

We performed several analyses to stress-test our *CACNA1C* instruments for thoracic aortic outcomes. First, we implemented a single-SNP model, which produced highly similar causal effect estimates for *CACNA1C* expression on TAA/D and thoracic aortic diameter (**Supplementary Figure 1**; **Supplementary Table 3**). Second, Steiger tests were applied, which indicated that our instrumental variables explained significantly more variance in *CACNA1C* expression than for TAA/D or thoracic aortic diameter (**Supplementary Table 7**), indicating absence of reverse causation. Third, we assessed for possible local genetic pleiotropy of our instruments. Reassuringly, none of the instruments, which were intronic to *CACNA1C*, were significant expression or splice QTLs for other genes in any tissue in the GTEx v10 dataset^11^. Together, these findings validate our *CACNA1C* instruments and Mendelian randomization framework.

Finally, given the complex interplay between *CACNA1C*, blood pressure, and thoracic aortic traits, we aimed to perform conditional/mediation analyses (**Supplementary Table 8**). We performed two-step Mendelian randomization to estimate causal effects of *CACNA1C* expression on TAA/D and ascending aortic diameter, conditional on the effect of *CACNA1C* expression on blood pressure (**Figure 1B**).

Consistent with the discordant effects of *CACNA1C* expression on aortic traits and on specific blood pressure traits (systolic and diastolic blood pressure), we observed stronger causal effects when accounting for these blood pressure traits in our two-step Mendelian randomization. For instance, when accounting for systolic blood pressure, the causal effect estimate of aortic *CACNA1C* expression became stronger on TAA/D (β=-0.19, 95% CI [−0.32, −0.07], *P*=0.002) and on ascending aortic diameter (β=-0.09, 95% CI [−0.14, −0.03], *P*=0.001) (**Figure 1E**). In contrast, accounting for pulse pressure weakened (but did not abolish) the estimated direct causal effect of *CACNA1C* expression on both aortic outcomes (**Figure 1E**).

Taken together, using human genetics, we provide evidence supporting an adverse effect of lower aortic expression of *CACNA1C*, the main aortic target of antihypertensive L-type CCBs, on thoracic aortic disease. Interestingly, the estimated causal effect of *CACNA1C* expression became stronger after accounting for systolic and diastolic blood pressure effects. These findings suggest that adverse aortic wall effects associated with L-type calcium channel blockade may be partially masked by beneficial blood-pressure-lowering effects. In contrast, the partial mediation effect observed for pulse pressure may indicate that changes in aortic wall stiffness^20–23^ - likely mediated through vascular smooth muscle cells^24–26^ - may explain some portion of the adverse effects of calcium channel blockade on TAA/D and aortic dimensions, although this potential mechanism requires further investigation.

When extrapolating our findings to L-type CCB medications, several considerations should be noted. First, drug-target Mendelian randomization may not fully recapitulate pharmacological intervention, as lifelong genetically-mediated differences in target expression levels may not reflect the magnitude, timing, tissue specificity, or local pharmacodynamics of actual drug exposure. Notably, however, our *CACNA1C* instruments recapitulated the expected effects of CCBs across various blood pressure traits, supporting the validity of our *CACNA1C* Mendelian randomization as a proxy for antihypertensive L-type CCBs. Second, Mendelian randomization is contingent on several assumptions, including the use of strong instruments and absence of pleiotropic pathways. To mitigate such issues, we restricted to *cis*-eQTLs^27^ with high F-statistics^28^, implemented Steiger directionality tests^29^, and found no evidence of local transcriptomic pleiotropy for our instruments. We also note that the strength of the statistical evidence of our MR results remains relatively modest, although we reassuringly found consistent results, across two different outcomes, that capture distinct components of thoracic aortic health and disease. Finally, although *CACNA1C* represents the primary pore-forming subunit of arterial calcium channels and represents the main target of L-type CCBs, our findings do not imply identical effects across all CCB subclasses and calcium channel subunits.

In summary, when viewed together with recent observational and experimental studies^6,30^, our results provide initial convergent evidence supporting a potential adverse effect of CCBs on thoracic aortic disease. The agreement between complementary lines of evidence strengthens the overall inference beyond any individual study design. While dedicated clinical trials remain necessary, these findings suggest that potential aortic effects should be considered when selecting antihypertensive therapy in patients with established TAA/D or elevated risk of thoracic aortic disease, particularly when alternative treatment options are available.

## Methods

### Genome-wide association study data sources for Mendelian randomization

We constructed a well-powered TAA/D GWAS by meta-analyzing multiple GWAS datasets from independent cohorts, including FinnGen^31^, UK Biobank^32^, CHIP/MGI^33^ and the Million Veteran Program (MVP)^18^. Details on the cohorts included in the TAA/D meta-analysis have been reported previously^12^. GWAS summary statistics of the independent cohorts were meta-analyzed using METAL with a fixed-effect inverse-variance weighted method after extensive quality control^12^. After meta-analysis, variants were filtered on MAF ≥1% to include common variants only. The TAA/D GWAS meta-analysis included data on 14,409 cases and 1,210,227 controls. Additionally, we used summary statistics for GWAS on ascending aortic diameter^9^. Genetic associations for systolic blood pressure, diastolic blood pressure and pulse pressure were obtained from a large GWAS meta-analysis including 1,028,980 individuals^34^. Hypertension, coronary artery disease and abdominal aortic aneurysm GWAS summary statistics were collected from the Cardiovascular Disease Knowledge Portal^35^. Variants were lifted to genome build GRCh38 with UCSC liftOver where necessary and formatted using MungeSumstats^36^ with dbSNP 155. Ambiguous variants, INDELs, variants with MAF <1% and imputation quality (INFO) score <0.6 (if available) were removed.

### Selection of eQTLs as genetic instruments

Genetic variants associated with aortic gene expression (*cis*-eQTLs) were obtained from GTEx v8^11^ (N=387 samples). For our main analysis, the instrumental variable for *CACNA1C* was defined by selecting variants from the LASSO model of the FUSION prediction weights^10^. Variants that were considered a strong instrument (*F*-statistic >10) and significant within the GTEx dataset were included. As a sensitivity analysis, we additionally constructed a *CACNA1C* instrumental variable with clumping. First, the GTEx *cis*-eQTL dataset was harmonized to respective outcome summary statistics and variants present in the LD reference, generated from 50,000 high-quality samples of European ancestry randomly selected from the UK Biobank. The instrumental variable was then defined by clumping *cis*-eQTLs in PLINK2^37^, using two-sided *P*<5×10^−5^, r^2^<5×10^−4^ and a 10 Mb window size. For both the multi-SNP model (derived from LASSO weights) and the single-SNP model (derived from clumping), additional Steiger filtering was performed to assess potential reverse causality^29^.

### Mendelian randomization analyses

We performed two-sample MR analyses to investigate the association between genetically predicted *CACNA1C* expression and TAA/D, ascending aortic diameter, and blood pressure traits. For the multi-SNP *CACNA1C* model, MR was performed using the MendelianRandomization R-package^38^ with the inverse-variance weighted (IVW) method. Linkage disequilibrium between variants included in the instrumental variable was accounted for using an LD correlation matrix generated with PLINK2. For the clumping-based single-SNP *CACNA1C* model, MR was performed using a Wald ratio test implemented in the TwoSampleMR R-package^39^.

We additionally performed two-sample MR to assess the association between blood pressure traits and aortic outcomes. Summary statistics were harmonized to include variants present in both datasets and restricted to those present in the LD reference, built from high-quality data of 5000 randomly selected participants of the UK Biobank. After harmonization and prior to MR, variants were filtered and pruned based on genome-wide significance (*P*<5×10^-8^) and r^2^<5×10^-4^, with a 10 Mb window size. The weighted median method implemented in TwoSampleMR was used, which is robust to outliers^17^.

### Two-step Mendelian randomization and mediation

To adjust the CACNA1C – aortic outcome associations for a potential mediation effect of blood pressure traits, we applied a two-step (network) MR approach^40^. We performed univariable two-sample MR to identify the effect of *CACNA1C* aortic expression on both TAA/D and ascending aortic dimension (β_total_), the effect of *CACNA1C* aortic expression on blood pressure traits (β_1_), and the effect of blood pressure on TAA/D and ascending aortic diameter (β_2_). The direct effect of *CACNA1C* on aortic outcomes, mitigating the potential mediation of blood pressure, was then estimated as β_total_ - β_1_×β_2_. Standard errors and 95% confidence intervals were estimated using the delta method.

## Acknowledgements

We gratefully thank all research participants of the biobanks included in the original publications (FinnGen, UK Biobank, CHIP+MGI and MVP). N.G. received research support from the Dutch Heart Foundation (03-003-2024-0102). C.R.B. was supported by funding from the Pathfinder Cardiogenomics program of the European Innovation Council of the European Union (DCM-NEXT and Nav1.5-CARED projects). S.J.J. was supported by a VENI grant from The Netherlands Organisation for Health Research and Development. C.R.B. and S.J.J. were also supported through the KIC program funded by the Dutch Research Council and Dutch Heart Foundation (PRECISE project).

## Data availability

FinnGen and UK Biobank meta-analysis summary statistics are publicly available after requesting access to FinnGen services (https://www.finngen.fi/en/access_results). CHIP+MGI summary statistics can be accessed through the University of Michigan Center for Statistical Genetics (https://csg.sph.umich.edu/willer/public/TAA2021). MVP summary statistics are available through dbGAP under accession code phs001672.v14.p1 (https://www.ncbi.nlm.nih.gov/projects/gap/cgi-bin/study.cgi?study_id=phs001672.v14.p1). Summary statistics for blood pressure traits are available through the GWAS Catalog website data repository (https://www.ebi.ac.uk/gwas/) with data accession codes GCST90310294, GCST90310295 and GCST90310296 for SBP, DBP and PP, respectively. Summary statistics for ascending aortic diameter, hypertension, coronary artery disease, and abdominal aortic aneurysm were downloaded from the Broad Institute Cardiovascular Disease Knowledge Portal (https://cvd.hugeamp.org). Aortic *cis*-eQTLs were obtained from GTEx v8 (https://www.gtexportal.org/home/downloads/adult-gtex/overview).

## Ethics approval

All datasets used in this study are publicly available, and included studies were conducted in accordance with relevant regulations. Ethics approvals were given for each study by institutional review boards and ethic committees, and participant enrollment, data collection, and data analysis complies with ethical regulations^9,11,18,31–35^.

