## Supplementary Figure 1 for "Genetic support for an adverse effect of calcium channel blockers on the human thoracic aorta"

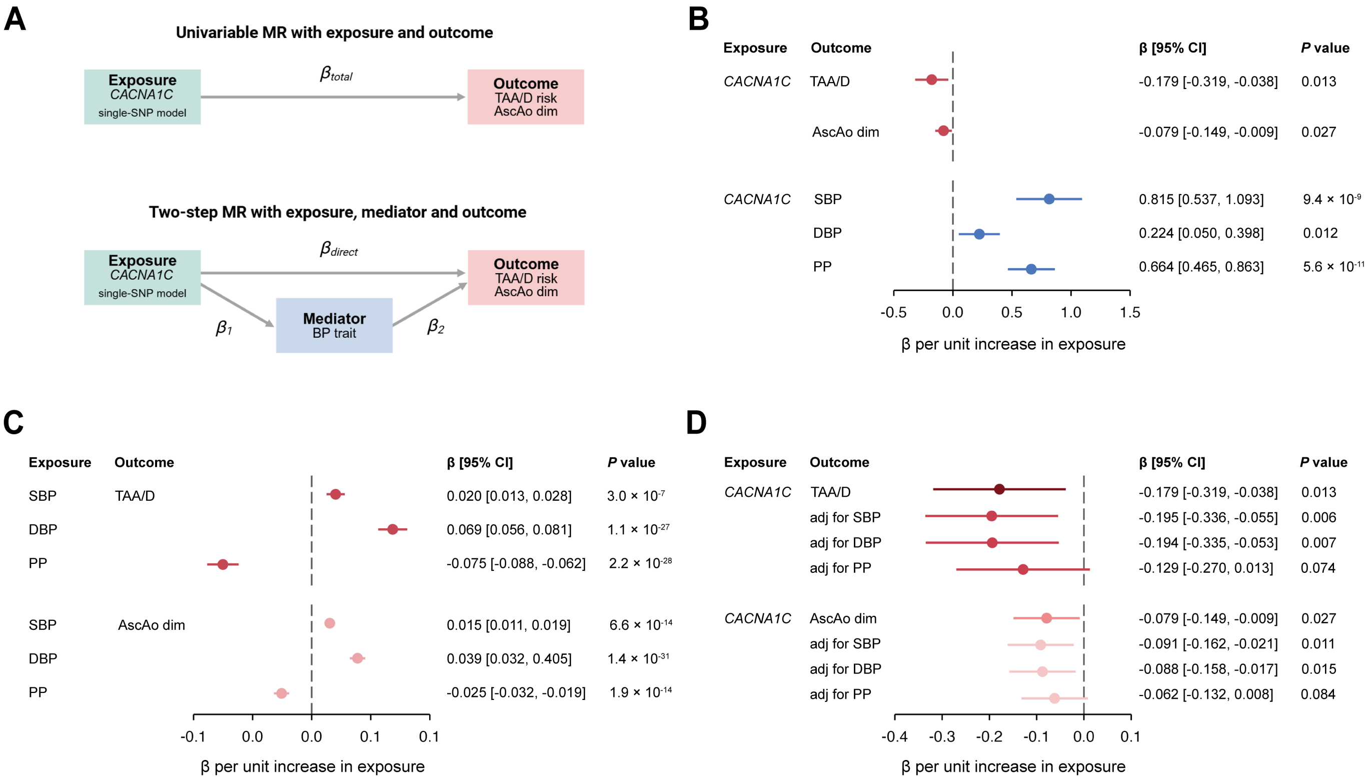


**Supplementary Figure 1 – Mendelian randomization analyses for aortic *CACNA1C* expression on aortic outcomes with a single-SNP model.** A, MR analyses to assess the effect of aortic *CACNA1C* expression on TAA/D risk and ascending aortic diameter with a single-SNP model. A potential mediation effect of blood pressure traits is accounted for using a two-step MR approach. B, Forest plot showing the results for univariable MR of aortic *CACNA1C* expression on both aortic outcomes and blood pressure traits for a single-SNP model. C, Forest plot showing the results of univariable MR for the effect of blood pressure traits on both aortic outcomes for a single-SNP model. D, Forest plot showing the results of two-step MR for the effect of aortic *CACNA1C* expression on aortic outcomes for a single-SNP model, adjusting for a potential mediation effect of blood pressure on both TAA/D risk and ascending aortic diameter. Note: MR, Mendelian randomization; TAA/D, thoracic aortic aneurysm and dissection; AscAo dim, ascending aortic diameter; SBP, systolic blood pressure; DBP, diastolic blood pressure, PP, pulse pressure.
